# Piloting a data-extraction form for verbatim terminology: a blinded inter-reviewer agreement study before a surgical evidence map

**DOI:** 10.64898/2026.09.18.26363356

**Authors:** Paulo Ayroza Ribeiro, Ana Clara Servidoni, Marina Paula Andres, Henrique Abrão, Guilherme Karam, Helizabet Salomão Abdalla Ayroza Ribeiro, Mauricio S. Abrão

## Abstract

**Background:** Data-extraction forms are tested before use in systematic reviews, but the guidance and error studies behind that practice concern numerical data. When the datum is a term transcribed verbatim from a source, as in evidence maps that inventory terminology, neither an agreement measure nor extraction conventions are established. We piloted, before Phase 1 of the ATLAS-ONTO project, a form built to extract operative-step terms from the surgical literature on deep endometriosis, to determine whether it produced acceptable inter-reviewer agreement and what revisions it required.

**Methods:** Blinded inter-reviewer agreement study, reported according to GRRAS. Two reviewers independently extracted terms from 11 purposively heterogeneous articles in English and French. The primary measure was the mean per-article Jaccard index of normalized term sets, with an explicit rule for empty sets; secondary measures were Cohen’s κ for categorical attributes of exactly matching terms and simple agreement for anatomical structure. Thresholds were fixed before extraction (Jaccard ≥0.70; κ ≥0.60; simple agreement ≥0.80). Unmatched terms were classified retrospectively by mechanism, and the same articles were reassessed after a harmonization session.

**Results:** Eight articles were extracted, seven with a defined index. The form failed its threshold: the mean Jaccard index was 0.581 (pooled 55/92 = 0.598). Of 92 terms, 55 matched exactly and 37 did not; the unmatched terms were attributed to source coverage (21), span extent (8), source-language comprehension (7) and one residual discrepancy. κ conditional on the 55 matching terms was high (end definition 0.930; laterality 0.781) and anatomical-structure agreement was 0.873, so κ alone did not detect the failure. Two record-integrity defects found during analysis are reported with their effect. Four written conventions and a split start-definition field followed; reassessment after harmonization reached 0.913, which is convergence, not independent reproducibility.

**Conclusions:** A form that extracts terminology or verbatim text should be piloted with a set-overlap measure, a rule for empty sets and a defined admissible source, against a threshold fixed in advance. The mechanisms of discordance found here are properties of text extraction and are not specific to surgery. The revised form will be tested on new material in Phase 1.

## 1. Background

Data extraction is the step of a systematic review at which the published record becomes data, and the guidance on it is consistent: extract in duplicate, pilot the form, and document the decisions [1,2]. The empirical basis for that guidance comes from studies of numerical extraction — effect estimates, sample sizes, outcome values — in which errors were frequent and duplicate extraction reduced them [3–5]. Those studies define error against a verifiable value in the source.

Terminology extraction has no such value. When the datum is the term an author used for an action — the object of evidence maps that inventory a field’s vocabulary — the reviewer decides what qualifies as a term, where its text span begins and ends, and which components of the source (text, tables, captions, video) count as evidence. Preserving the original wording is necessary to study lexical variation, but exact-string disagreement may then reflect different spans describing the same action rather than different concepts. Whether two reviewers extract the same terms from the same article is therefore an empirical question that the numerical error literature does not answer, and for which the usual coefficient, Cohen’s κ, is ill suited: κ needs paired observations, and unmatched terms have no pair.

The case here is the surgical literature on deep endometriosis. Existing frameworks describe related but distinct levels — hysteroscopic terminology [6], imaging-based descriptions [7], a visual lesion ontology [8], the anatomy-based surgical complexity classification of the AAGL [9], the international endometriosis terminology [10] and the SAGES surgical-gesture taxonomy [11] — and none inventories the operative steps named in the literature. ATLAS-ONTO will derive that inventory by systematic evidence mapping and submit it to international consensus.

Before the definitive extraction, the form was tested. This paper reports that pre-Phase 1 pilot as a reliability and agreement study of the instrument: whether two reviewers, blind to each other, extracted the same terms against thresholds fixed in advance; what the discordance consisted of; and what was changed as a result. It evaluates the form, not the ontology or its clinical validity, and it is written for readers who extract terminology or verbatim text in any field.

## 2. Methods

### 2.1 Design, terminology and reporting

This pre-Phase 1 methodological pilot is reported according to the Guidelines for Reporting Reliability and Agreement Studies (GRRAS) [12]; the Phase 1 systematic review and the later Delphi consensus are not reported here. The pilot was conducted under the parent protocol version 2.3 (17 August 2026) and the analysis plan ATLAS-EST-02 version 1.3, in which the measures, thresholds and failure rule were fixed before extraction began. The Open Science Framework registration (https://doi.org/10.17605/OSF.IO/GMXZ5) was created on 4 September 2026, after the pilot, so that it would hold the registered protocol and analysis plan (versions 2.9 and 2.2), the datasets of every round, the scripts and the complete outputs; it time-stamps the deposit, not the fixing of the thresholds (section 4.2).

We distinguish blinded inter-reviewer agreement from post-harmonization convergence on previously discussed material. Reproducibility of the revised form on new material with independent reviewers was not assessed.

The principal measurement was exact term-set overlap, summarized as the mean of the per-article Jaccard indices over the articles with a defined index, as specified in the plan. Attribute agreement was secondary and conditional on terms matching after normalization; it does not measure completeness or accuracy of the whole extraction.

### 2.2 Observers

Reviewer 1 was a gynecologic surgeon with 30 years of surgical experience and limited systematic-review experience; Reviewer 2 was a medical student with no surgical experience and extensive systematic-review experience. Declared reading languages were English, Spanish and French. Both received the manual and a worked example before the blinded round; neither had access to the preliminary ontology, and neither used a translation aid in the blinded round.

### 2.3 Sample

Eleven articles were purposively selected by the steering committee to challenge the instrument across English- and French-language surgical videos, technique descriptions, clinical series, classifications, computational research and a practice survey (Table 1) [8,9,13–21]; some concerned adjacent gynecologic procedures. It was a stress-test sample, not a representative one. Three articles were expected to yield few or no terms; two full texts were not retrieved at the time of extraction.

**Table 1.**
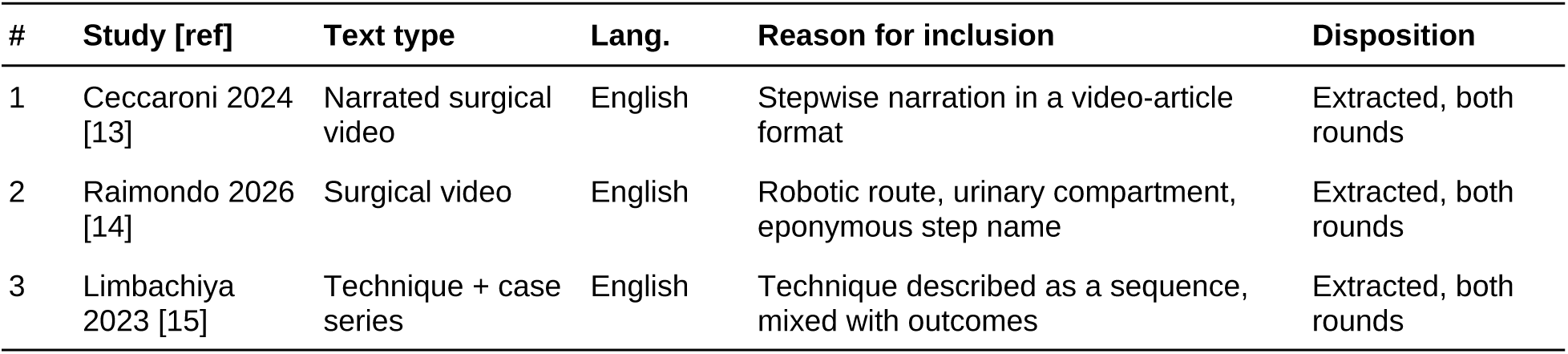
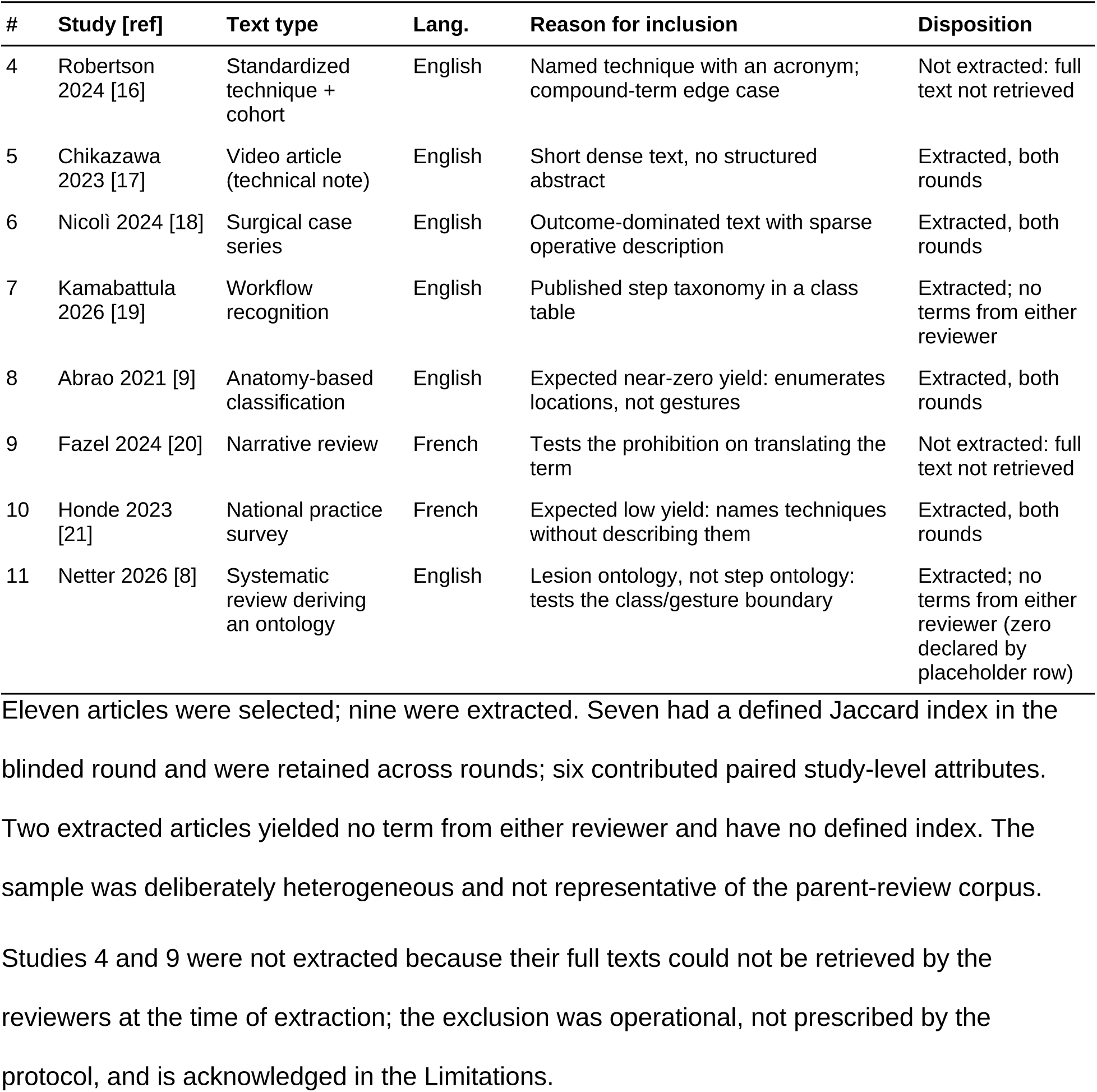
Purposive stress-testing sample and extraction status.

### 2.4 The extraction form

Each row recorded one extracted term, verbatim in the original language, with its language, anatomical structure, laterality, presence of an operational start and end definition, and supporting quotations. Procedure class and access route were study-level fields. An article yielding no term was declared with a single placeholder row. No mapping to the preliminary ontology was permitted. The blank form and the manual, initial and revised, are Supplementary Materials S1 and S2.

### 2.5 Phases

The reviewers received the manual on 18 August 2026 and extracted independently between 18 and 31 August, each in a separate, unshared copy of the form (blinded round). After the threshold was not met, discordant records were discussed in a harmonization session on 31 August–1 September and three written conventions were adopted; a fourth, on source language, followed. Reviewer 1 then revised her records on the same articles while Reviewer 2’s were unchanged, so the post-harmonization round is convergence after discussion, not blinded re-extraction. Two intervening exploratory exercises compared an LLM-generated extraction audited by Reviewer 1 with Reviewer 2’s records; their outputs are excluded from all estimates but informed the conventions (Supplementary Table S6).

### 2.6 Statistical analysis

Analyses were descriptive; no hypothesis was tested and no p value is reported. The thresholds fixed in the analysis plan were a mean study-level Jaccard index ≥0.70, Cohen’s κ ≥0.60 for each closed-list field and simple agreement ≥0.80 for anatomical structure — decision rules for the instrument, not validated universal standards [2,22]. The failure rule required retraining, revision of the form and repeat testing; repeat testing on independent material was deferred to Phase 1 (section 2.7).

For each article, the Jaccard index was the size of the intersection of the two reviewers’ normalized term sets divided by the size of their union. A set empty for one reviewer only gives zero; two empty sets give an undefined value (0/0), and such articles leave the mean with their number reported, as prespecified — the empty–empty case is assigned neither zero nor one. The primary estimate is the arithmetic mean of the per-article indices over the articles with a defined index; it is sensitive to articles with few terms, where one disagreement moves the index a long way, so the pooled index (sum of intersections over sum of unions) and the mean over the articles with terms from both reviewers are reported alongside. Anatomical-structure agreement required identity of the declared sets after splitting on commas and semicolons and normalization.

Normalization was deliberately minimal: case folding, trimming of outer whitespace and collapse of internal whitespace, and nothing else. Accents, hyphens, punctuation, stop words, stemming, translation and synonym mapping were left untouched, because terminological variation is the object of the parent review and any mapping would remove the phenomenon the review exists to measure. Within-article repetitions retained the first occurrence: of 150 term rows, three were collapsed, leaving 147 reviewer-specific records across seven articles (59 for Reviewer 1, 88 for Reviewer 2). One further row was a placeholder declaring a zero-term article, not a term (section 3.1).

Term attributes were paired only through exact normalized matches — the intersection that is the numerator of the Jaccard index — with no matching by similarity, string distance or human judgement, since semantic similarity is what the parent review studies and cannot enter the measure that evaluates it. κ therefore acts on the terms that both reviewers found and transcribed identically; unmatched terms contribute nothing to it, so high conditional agreement cannot establish overall extraction performance, and the Jaccard index is the counterweight.

Procedure-class and access-route coefficients used the article, not the term row, as the unit, because those are attributes of the article recorded on a sheet whose unit is the term, and computing them per term would count one agreement many times.

Cohen’s κ is reported with observed agreement and marginal counts [23,24]. The 95% confidence intervals use a normal approximation truncated to the parameter range; they are nominal and ignore clustering of terms within articles. Where both reviewers used a single, identical category, κ is not estimable and is reported as such with observed agreement; where one reviewer alone used a single category, κ equals zero by construction despite non-zero observed agreement. No confidence interval was calculated for the Jaccard index.

### 2.7 Planned external verification

The parent protocol prespecifies external re-assessment of 20% of screening records and of extractions by a third reviewer who took part in neither the original extraction nor the adjudication, on the definitive corpus and under the conventions in force, with sampling frames, comparator and escalation rules fixed in the registered analysis plan. Three questions are kept apart in that plan and here: agreement between the two original reviewers before adjudication, which is the only measure between independent observers; agreement of the third reviewer with the adjudicated dataset, which is agreement with a reference, not accuracy against an established truth; and reproducibility of the revised instrument with different reviewers on different material, which this pilot does not measure and declares as a limitation.

### 2.8 Software and analytical reproducibility

Analyses used base R 4.3.3 (R Foundation for Statistical Computing, Vienna, Austria), with every coefficient implemented from its definition and independently recomputed in Python 3.11.15 with scikit-learn 1.8.0; all values agreed to three decimal places. Two record-integrity defects found while the results were regenerated on 2 September 2026 are reported in Supplementary Table S9: a study-name matching defect under a non-UTF-8 locale that silently omitted Nicolì et al [18]. (mean inflated to 0.639), and a placeholder row declaring a zero-term article [8] that had been counted as a term (eighth index of zero; mean 0.508). All values reported here were regenerated after both corrections. Raw datasets, scripts, execution logs and cross-check outputs are deposited in the registration; the corrected script (version 1.1) and its output, produced after the registration, are in the associated public project (see Data availability).

### 2.9 Ethics

The study analyzed published articles and the extraction records of two of its authors, acting as reviewers. No patient, human research participant or personal data outside the authorship were involved, and ethics-committee review was therefore not sought. Both reviewers consented to the reporting of their records and professional profiles.

### 2.10 Use of large language models

Large language models were used for four distinct purposes, none of which produced any estimate reported here: (1) Claude (Anthropic; Claude Opus 4.8 in August 2026 and Claude Fable 5.1 from September 2026, accessed through the Claude desktop application) to identify candidate articles for the purposive sample, each verified by the authors against the primary record; (2) Claude (Claude Opus 4.8, same access, 31 August–1 September 2026) to generate the exploratory pre-extraction and the post-blinded-round appraisal that Reviewer 1 audited (section 2.5; Supplementary Table S3), whose outputs are excluded from all estimates but informed the conventions; (3) Claude (same models and access, August–September 2026) to assist with the analysis scripts, every coefficient of which was implemented from its definition, checked line by line and independently recomputed in Python; (4) Claude and ChatGPT (OpenAI; GPT-5, 4 September 2026) to assist with drafting, methodological review and language editing. The blinded-round and post-harmonization extraction records were entered by the two reviewers without AI assistance. The authors reviewed and edited all content and take full responsibility for it. No AI tool is an author.

## 3. Results

### 3.1 Studies extracted

Nine of the 11 selected articles were extracted; two full texts were not retrieved at the time of extraction [16,20]. Two extracted articles yielded no term from either reviewer: Kamabattula et al. [19], for which neither reviewer entered a row, and Netter et al. [8], for which Reviewer 1 entered a placeholder row declaring zero terms. Seven articles therefore had a defined blinded-round Jaccard index and were retained across rounds; six contributed paired study-level attributes (Table 1).

An LLM-assisted, reviewer-audited appraisal performed after the blinded round classified the nine articles’ suitability as terminology sources (Supplementary Table S3). It was neither blinded nor an independent reference standard.

### 3.2 Agreement in the blinded round

The mean Jaccard index was 0.581 across the seven articles with a defined index (pooled 55/92 = 0.598) and 0.677 across the six with terms from both reviewers. Neither met 0.70. The seven articles yielded 59 and 88 reviewer-specific terms, with 55 exact matches (Table 2).

**Table 2.**
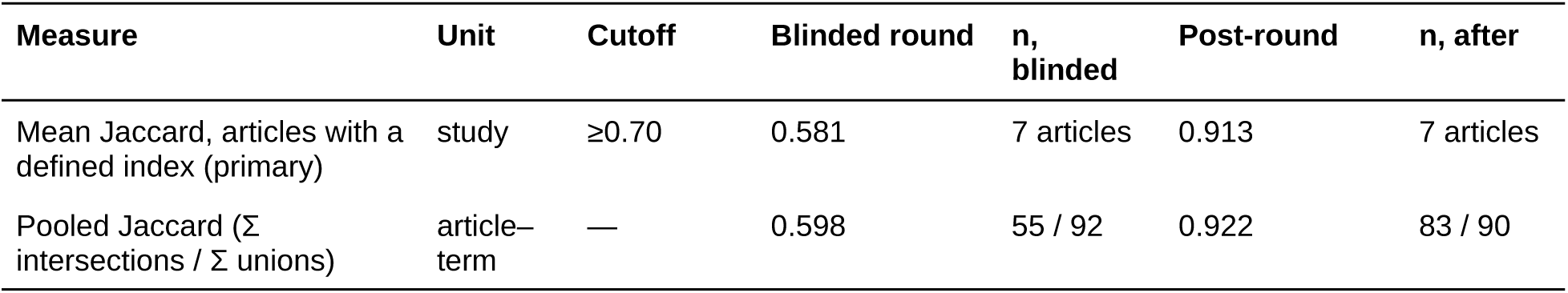
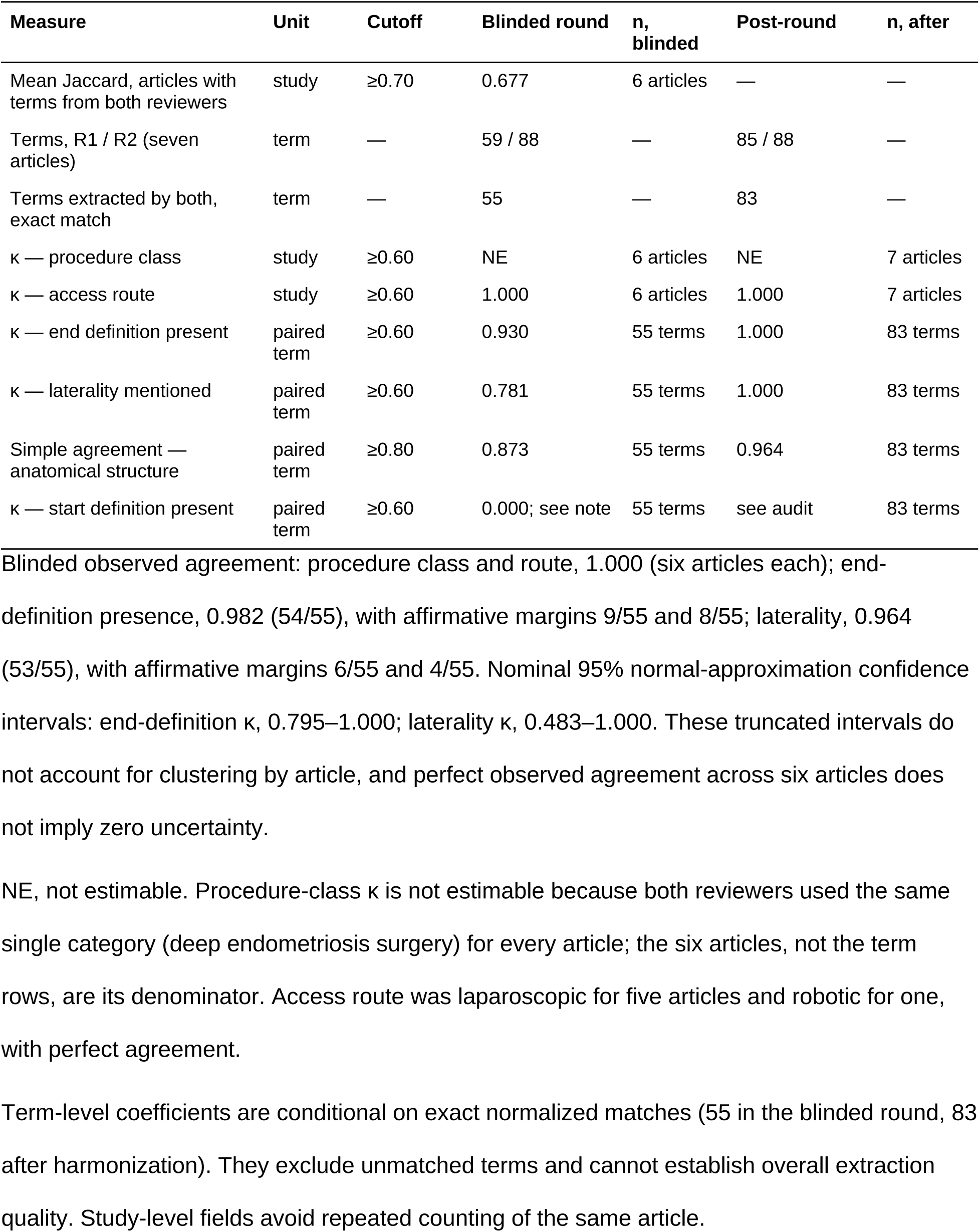
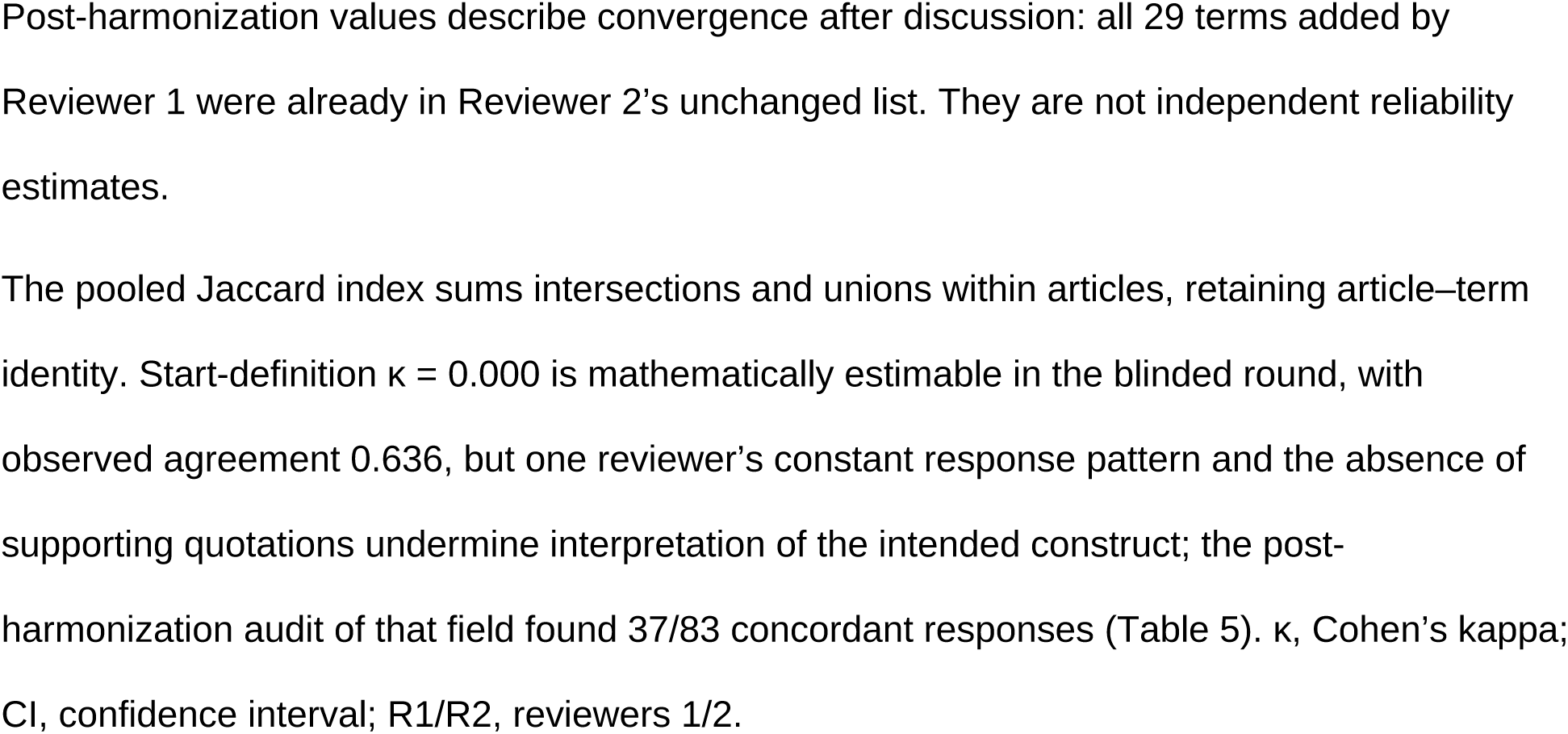
Blinded-round agreement and post-harmonization convergence, with analysis units and denominators.

Among the 55 matched terms, end-definition κ was 0.930 (95% CI 0.795–1.000; observed agreement 0.982) and laterality κ 0.781 (95% CI 0.483–1.000; observed agreement 0.964), resting on one and two discordant pairs respectively; anatomical-structure agreement was 0.873. Across the six articles with paired study-level attributes, access-route κ was 1.000 and procedure-class κ was not estimable because a single category occurred. The laterality interval includes values below the threshold; the start-definition field is reported separately below.

### 3.3 Retrospective characterization of discordance

The seven-article union contained 92 terms: 55 matching and 37 unmatched. Retrospective review attributed 21 unmatched terms to source coverage, eight to span extent, seven to source-language comprehension and one to a residual discrepancy (Table 3). These attributions are explanatory judgments made after the fact, not experimentally established causes.

**Table 3.**
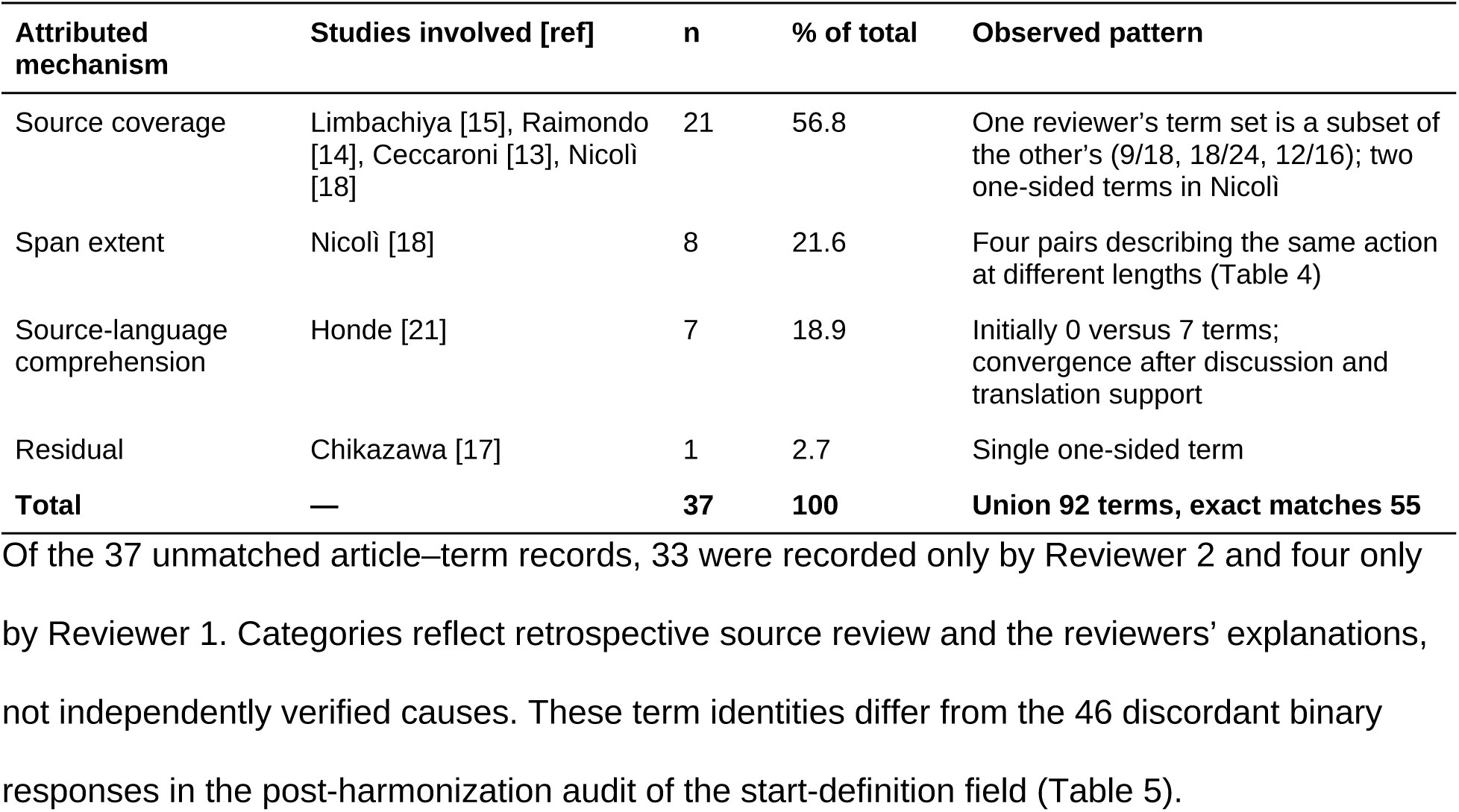
Retrospective classification of 37 unmatched terms in the seven-article blinded-round subset.

| Attributed mechanism | Studies involved [ref] | n | % of total | Observed pattern |
| --- | --- | --- | --- | --- |
| Source coverage | Limbachiya [15], Raimondo [14], Ceccaroni [13], Nicoli [18] | 21 | 56.8 | One reviewer's term set is a subset of the other's (9/18, 18/24, 12/16); two one-sided terms in Nicoli |
| Span extent | Nicoli [18] | 8 | 21.6 | Four pairs describing the same action at different lengths (Table 4) |
| Source-language comprehension | Honde [21] | 7 | 18.9 | Initially 0 versus 7 terms; convergence after discussion and translation support |
| Residual | Chikazawa [17] | 1 | 2.7 | Single one-sided term |
| **Total** | — | **37** | **100** | **Union 92 terms, exact matches 55** |

Coverage-related cases were articles in which one reviewer’s terms were a subset of the other’s [13–15]; nesting suggests different coverage or eligibility decisions but does not by itself distinguish them.

Four pairs of terms from Nicolì et al. [18], each identified by both reviewers but transcribed at different lengths, account for the eight span-related strings (Table 4). The two remaining unmatched terms in that article were recorded by Reviewer 2 only and are counted under source coverage.

**Table 4.** Span-extent discordance in Nicolì 2024 [18]: four steps transcribed at different lengths.

| Reviewer 1, as transcribed | Reviewer 2, as transcribed |
| --- | --- |
| skin incision | 2-cm skin incision |
| insufflation CO2 | insufflation CO2 into the intraperitoneal cavity |
| pronated arms should be positioned | pronated arms should be positioned along the body or form an angle <90° with the operating table |
| Foley catheter is inserted into the bladder | 14/18 G Foley catheter is inserted into the bladder and a RUMI manipulator |

In the French survey [21], Reviewer 1 initially extracted no terms; after discussion and use of a translation for comprehension she recorded the same seven original-language terms as Reviewer 2. Language support and harmonization effects cannot be separated.

### 3.4 Diagnostic audit of the start-definition field

Blinded-round start-definition κ was 0.000 with observed agreement 0.636 among the 55 matched terms: Reviewer 2 answered “yes” in all 90 of her original rows, and supporting quotations were absent in 40/40 affirmative rows for Reviewer 1 and 85/90 for Reviewer 2. The coefficient describes the recorded responses, not whether either reviewer applied the intended criterion.

A post-hoc audit then examined the 83 matched terms after harmonization — a different population from the 37 unmatched blinded-round terms. Responses disagreed in 46/83 pairs, 45 in the same direction, and 39 occurred in the three video articles (Table 5). Because these records followed discussion, this is not independent confirmation.

**Table 5.**
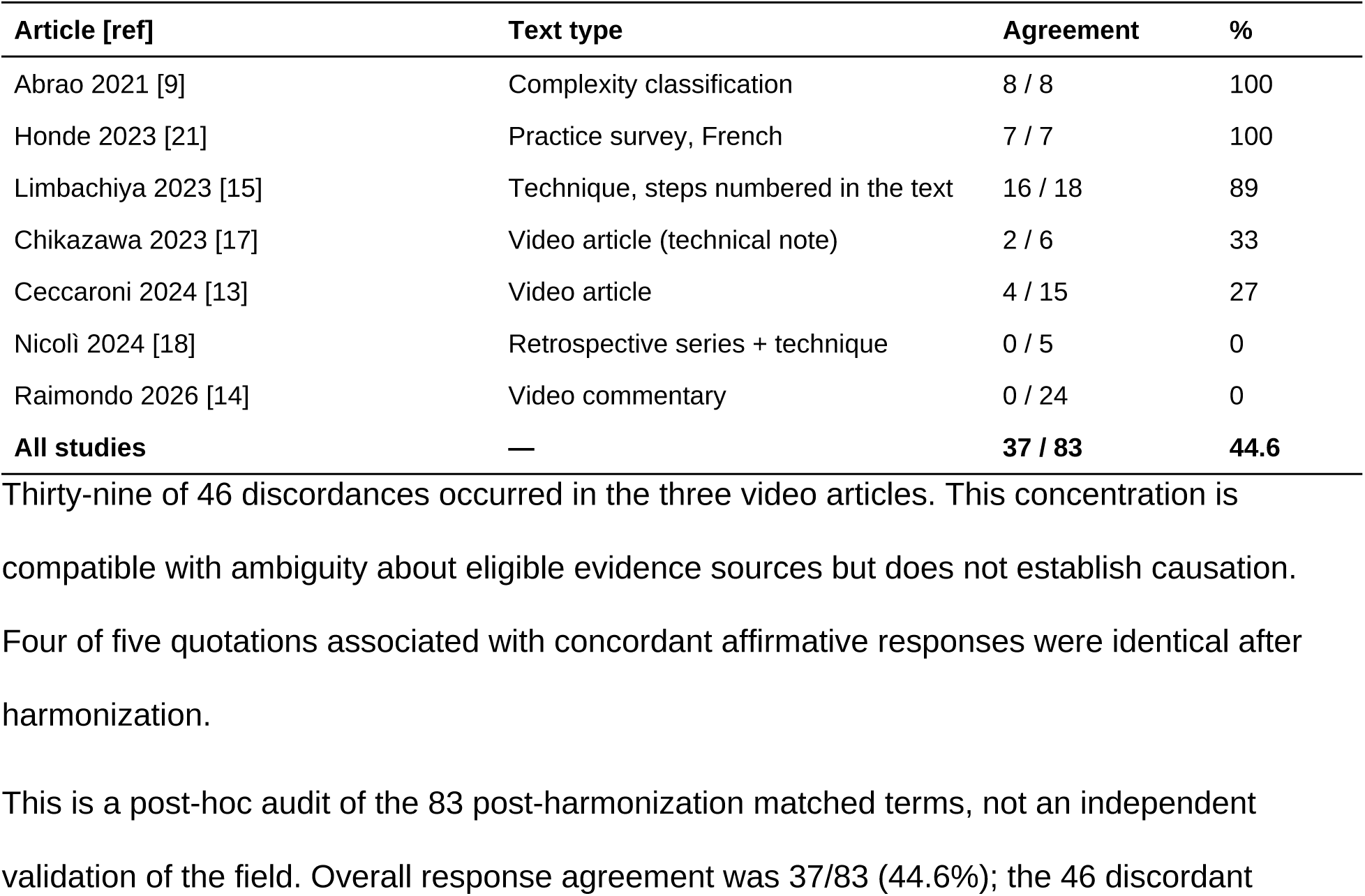
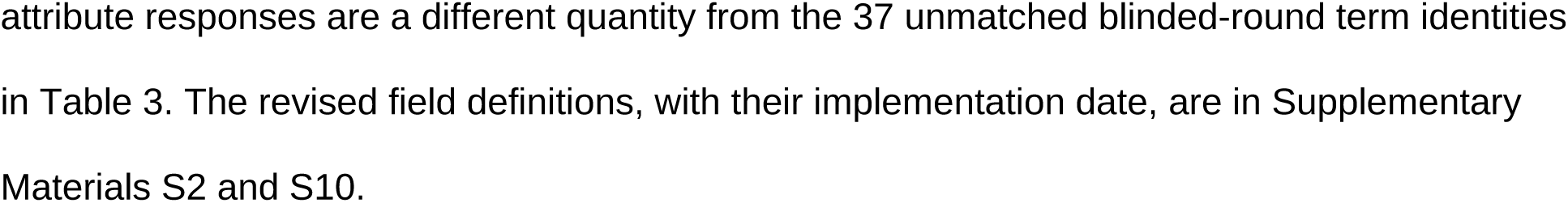
Post-hoc diagnostic audit of the original start-definition field, by article and text type.

The pattern suggested confusion between a step demonstrated on video and a start definition stated in the text. It does not establish the prevalence of missing definitions in the literature. The field was split into a text-definition field and a separate video-demonstration field, with a mandatory quotation (S2, S10).

### 3.5 The round after harmonization

After harmonization, the seven-article mean Jaccard was 0.913 (pooled 83/90 = 0.922); anatomical agreement was 0.964. Access-route, end-definition, and laterality κ values were 1.000, but procedure remained non-estimable and the start-definition field remained problematic. Nicolì et al [18]. remained below 0.70 (Tables 2 and 6; Fig. 1).

**Fig. 1.**
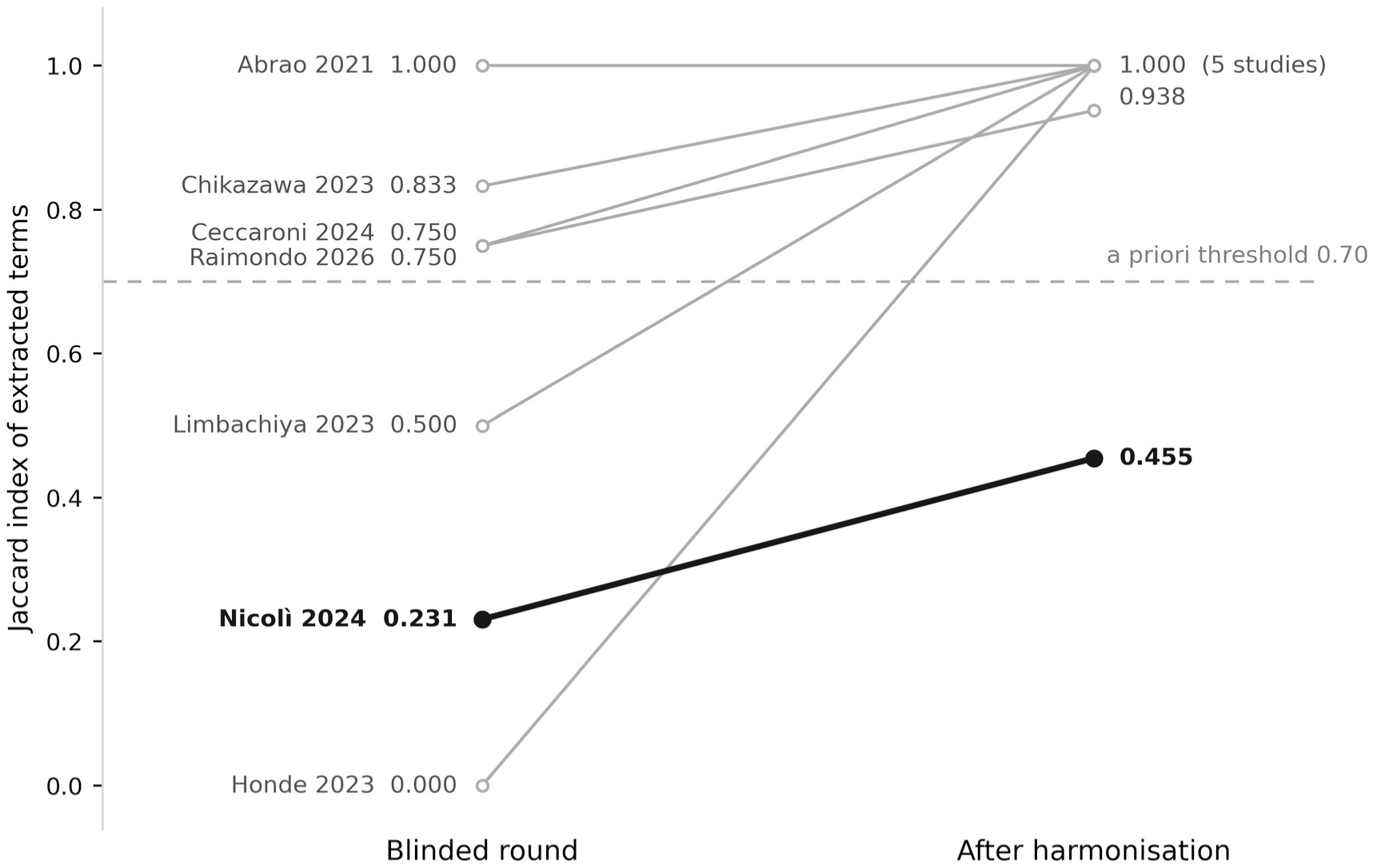
Per-article Jaccard index in the blinded round and after harmonization. Jaccard index of extracted terms for each of the seven articles with a defined index, in the blinded round and in the round after harmonization. The dashed line marks the prespecified threshold of 0.70. Nicolì 2024 [18] remains below the threshold after harmonization. The two extracted articles with no term from either reviewer have no defined index and are not shown. The post-harmonization mean of 0.913 summarizes convergence after discussion, not independent reproducibility.

**Table 6.** Jaccard index of extracted terms by study, blinded round and round after harmonization.

| Study [ref] | Blinded: R1 / R2 | Matches | Union | Blinded J | After J | $\Delta$ |
| --- | --- | --- | --- | --- | --- | --- |
| Abrao 2021 [9] | 8 / 8 | 8 | 8 | 1.000 | 1.000 | 0.000 |
| Ceccaroni 2024 [13] | 12 / 16 | 12 | 16 | 0.750 | 0.938 | +0.188 |
| Chikazawa 2023 [17] | 5 / 6 | 5 | 6 | 0.833 | 1.000 | +0.167 |
| Honde 2023 [21] | 0 / 7 | 0 | 7 | 0.000 | 1.000 | +1.000 |
| Limbachiya 2023 [15] | 9 / 18 | 9 | 18 | 0.500 | 1.000 | +0.500 |
| Nicoli 2024 [18] | 7 / 9 | 3 | 13 | 0.231 | 0.455 | +0.224 |
| Raimondo 2026 [14] | 18 / 24 | 18 | 24 | 0.750 | 1.000 | +0.250 |
| **Mean** | — | — | — | **0.581** | **0.913** | **+0.333** |

Kamabattula 2026 [19] and Netter 2026 [8] yielded no term from either reviewer and have no defined index. Nicoli 2024 [18] was the only article below 0.70 in both rounds; Honde 2023 [21] also failed initially ( $J = 0$ ). $\Delta$ is the change between rounds; the post-harmonization values are convergence after discussion, not an independent estimate.
R1: reviewer 1; R2: reviewer 2; J: Jaccard index; $\Delta$ : change in the Jaccard index between rounds.

Reviewer 1 retained 56 terms, removed three and added 29, reaching 85; all 29 additions were already in Reviewer 2’s unchanged 88-term list. This is convergence following discussion, not independent validation.

## 4. Discussion

The initial form did not meet the Jaccard threshold under any reported denominator. For a pre-Phase 1 study that result is informative rather than merely negative: it identifies the decisions a terminology-extraction manual must specify before scale-up, while preserving the distinction between original wording and interpreted concepts.

The observed patterns are compatible with instrument ambiguity, reviewer experience, language proficiency and heterogeneous source formats; this design cannot isolate their contributions.

A revised manual should define eligible actions and text spans, identify in-scope article/video components, and require a source locator for each record. Nested term sets warrant source-level checking, not an automatic diagnosis of incomplete reading.

Comprehension support should be distinguished from translating the extracted label. The original span should remain traceable; tool use and language proficiency should be documented, with unresolved passages referred for verification rather than coded as true absence.

Exact lexical overlap and agreement on attributes answer different questions. Neither supplies a gold standard for clinical concepts. Reporting both avoids interpreting high conditional κ or post-discussion convergence as evidence that the inventory is complete, correct, or reproducible.

The start-definition audit supports separating explicit textual evidence from video-demonstrated events. Whether such distinctions can be applied consistently requires fresh blinded testing, not further adjudication of the same records.

### 4.1 Implications for evidence synthesis

Three implications extend beyond the case. First, piloting an extraction form should be a measurement with a threshold fixed in advance, and the measure should fit the datum: for terms and verbatim spans, agreement is a question of set overlap, and a set measure such as the Jaccard index — with an explicit rule for empty sets — answers it, whereas κ on the paired subset answered a different question and was reassuring while the form failed. Second, the three mechanisms of discordance found here are properties of text extraction, not of surgery: reviewers read different parts of the source (coverage), delimit the same span differently (extent), and read a foreign-language source with unequal comprehension (language). Each is preventable by a written convention, and each is invisible to a κ computed on matched pairs.

Third, the start-definition audit shows what happens when the admissible source of an attribute is left undefined: one reviewer read a step demonstrated on video as a definition and the other did not, and the field measured reviewer interpretation rather than the literature. Forms that extract from multimodal sources should state which modality counts.

For the surgical field, a reproducible operative-step vocabulary could support reporting, training, video annotation and multicenter research; existing disease terminology and gesture frameworks should be reused where appropriate [10,11]. This pilot establishes neither a preferred technique nor improved outcomes; its contribution to that field is a tested instrument and a visible pathway to consensus.

### 4.2 Limitations

The small purposive sample and two reviewers preclude population-level reliability estimates. Expertise and language proficiency differed. One French article was extracted and another was unavailable; interpretation of the language effect is therefore limited.

There was no independent test of the revised form. Discussion, training, translation, record editing, and intervening LLM-assisted exercises were entangled, preventing attribution of convergence to any single change.

Analysis populations differ across outcomes by design (seven articles, six articles, 55 paired terms) and each estimate carries its denominator; conditional attribute analyses omit unmatched terms, and nominal term-level intervals ignore article-level clustering. The registration postdates the pilot and time-stamps the deposit; the claim that the thresholds preceded extraction rests on the dated plan and protocol cited in section 2.1.

Two unavailable articles reduced the intended range of text types. Retrospective disagreement coding and appraisal are vulnerable to confirmation bias, no independent reference standard was available, and the pilot cannot establish how standardized the wider literature is.

### 4.3 What was changed

Revisions addressed step recognition, span extent, source coverage, language support and the evidence required for a start definition; the four conventions and the split field are in the revised manual with a dated change log (S2, S10). The revised form and manual are frozen before Phase 1; the definitive extraction uses those versioned materials, and the nine pilot articles are re-extracted under them, so that no pilot record enters the parent inventory.

## 5. Conclusions

A data-extraction form for verbatim terminology failed a blinded pilot against a threshold fixed in advance, and the failure identified the decisions a terminology-extraction manual must specify: what counts as a term, how far its span extends, which parts of the source are read, how a source in another language is handled, and what counts as evidence for an attribute. Convergence after discussion is encouraging but does not demonstrate independent reproducibility; reliability claims require blinded verification on new material. The procedure — set-overlap measure, prespecified threshold, empty-set rule and written conventions — is available to any review that extracts terms or text rather than numbers.

## Supporting information

https://www.dropbox.com/scl/fi/h26nloyum7d4l8dpkw6et/Additional_file_1_Supplementary_Material_S3-S10.pdf?rlkey=9pznxqsu5gd4sb1t0m6pwratm&dl=0

## Data Availability

All data referred to in the manuscript are publicly available. The datasets supporting the conclusions of this article are deposited in the Open Science Framework registration of the study (https://doi.org/10.17605/OSF.IO/GMXZ5): the blank extraction forms (initial and revised), the extraction manual with its dated change log, the datasets of every round of the pilot (blinded round, the two exploratory exercises, the post-harmonization round, the corrected dataset and the directed audit), the audited pre-extraction and appraisal files, the data dictionary, the analysis scripts with the R version and locale guard, the Python cross-check, the consolidated results file and the complete analytical outputs before and after the two corrections described in the Methods. The corrected analysis script (version 1.1) and its output, produced after the registration and reproducing every estimate reported in the manuscript, are in the associated public OSF project (https://osf.io/vca2s/, folder "Post-registration corrections"). No individual patient or participant data were used.

## List of abbreviations

AAGL: American Association of Gynecologic Laparoscopists
CI: confidence interval
GRRAS: Guidelines for Reporting Reliability and Agreement Studies
J: Jaccard index
LLM: large language model
OSF: Open Science Framework
Po: observed proportion of agreement
R1, R2: Reviewer 1, Reviewer 2
SAGES: Society of American Gastrointestinal and Endoscopic Surgeons
Κ: Cohen’s kappa

## Declarations

### Ethics approval and consent to participate

The study analyzed published articles and the extraction records of two of its authors, acting as reviewers. No patient, human research participant or personal data outside the authorship were involved; ethics-committee review was therefore not sought. Both reviewers consented to the reporting of their records and professional profiles.

### Consent for publication

Not applicable. No data or images of individual persons are reported. The two reviewers, who are authors of this article, consented to the reporting of their extraction records and professional profiles.

### Availability of data and materials

The datasets supporting the conclusions of this article are available in the Open Science Framework registration of the study, https://doi.org/10.17605/OSF.IO/GMXZ5 — the blank forms (initial and revised), the extraction manual with its dated change log, the datasets of every round of the pilot (blinded round, the two exploratory exercises, the post-harmonization round, the corrected dataset and the directed audit), the audited pre-extraction and appraisal files, the data dictionary, the analysis scripts with the R version and locale guard, the Python cross-check, the consolidated results file and the complete analytical outputs before and after the two corrections described in section 2.8 — and in the associated public project, https://osf.io/vca2s/ (folder “Post-registration corrections”), which holds the corrected analysis script (version 1.1) and its output, produced after the registration and reproducing every estimate reported here.

### Authors’ contributions

Paulo Ayroza Ribeiro: Conceptualization, Methodology, Project administration, Supervision, Writing – original draft, Writing – review & editing. Ana Clara Servidoni (Reviewer 2): Investigation, Data curation, Formal analysis, Visualization, Writing – review & editing. Marina Paula Andres: Validation, Formal analysis, Writing – review & editing. Henrique Abrão: Writing – original draft, Writing – review & editing. Guilherme Karam: Validation, Writing – review & editing. Helizabet Salomão Abdalla Ayroza Ribeiro (Reviewer 1): Investigation, Data curation, Formal analysis, Writing – original draft, Writing – review & editing. Mauricio S. Abrão: Formal analysis, Validation, Writing – original draft, Writing – review & editing. All authors read and approved the final manuscript and agree to be accountable for all aspects of the work.

## Acknowledgements

Not applicable.

## Additional files

**Additional file 1:** Supplementary Material S3, S4, S6, S7, S8, S9 and S10 (PDF). S3, post-blinded-round exploratory appraisal of the nine extracted articles, with its provenance; S4, governance — competing interests, article-specific adjudication recusals and their record; S6, chronology of the pilot and data provenance (Table S6); S7, full contingency tables for every κ; S8, completed GRRAS checklist with manuscript locations; S9, computational integrity audit of the two record-integrity defects (Table S9); S10, the four extraction conventions and the evidence-traceability rules. In the text, “Supplementary Material S…” and “Supplementary Table S…” refer to the sections of this file.

**Additional file 2:** S1, blank extraction form, initial version as used in the blinded round (XLSX). The revised form (Appendix B v3.0), the extraction manual with its change log (S2, Appendix B-1 v3.0) and the data dictionary (S5) are deposited in the registration, https://doi.org/10.17605/OSF.IO/GMXZ5.

