## Supplementary material for "Piloting a data-extraction form for verbatim terminology: a blinded inter-reviewer agreement study before a surgical evidence map": https://www.dropbox.com/scl/fi/h26nloyum7d4l8dpkw6et/Additional_file_1_Supplementary_Material_S3-S10.pdf?rlkey=9pznxqsu5gd4sb1t0m6pwratm&dl=0

**Additional file 1 — Supplementary Material S3, S4, S6–S10**

Additional file to the article submitted to Systematic Reviews (Methodology). Not copy-edited. Reference numbers in square brackets follow the main article.

**Contents**

S3. Post-blinded-round exploratory appraisal of the nine extracted articles

S4. Governance: competing interests and adjudication recusals

S6. Chronology of the pilot and data provenance (Table S6)

S7. Full contingency tables

S8. GRRAS checklist

S9. Computational integrity audit of the two record-integrity defects (Table S9)

S10. The four extraction conventions and the evidence-traceability rules

Deposited separately: S1 (initial extraction form, Supplementary File S1; revised form, Appendix B v3.0, in the registration), S2 (extraction manual and change log, in the registration), S5 (data dictionary, in the registration). Registration: https://doi.org/10.17605/OSF.IO/GMXZ5. Associated public project with the corrected analysis script (v1.1) and its output: https://osf.io/vca2s/

**S3. Post-blinded-round exploratory appraisal of the nine extracted articles**

Provenance. An LLM generated this exploratory appraisal after the blinded round; Reviewer 1 audited it with access to the prior extractions. It is a nonblinded descriptive exercise, not pre-extraction eligibility screening or an independent reference standard.

| **Judgment as a source of operative steps** | **Studies, n of 9** |
| --- | --- |
| Full source — beginning, middle and end of the step described | 1 |
| Partial source — included with partial data | 3 |
| Methodological source, containing no operative step | 1 |
| Not a source of operative steps | 4 |

**Named-step counts in the exploratory appraisal**

| **Study [ref]** | **Named steps, n** |
| --- | --- |
| Abrao 2021 [9] | 11 |
| Limbachiya 2023 [15] | 10 |
| Chikazawa 2023 [17] | 5 |
| Raimondo 2026 [14] | 3 |
| Nicolì 2024 [18] | 2 |
| Ceccaroni 2024 [13] | 0 |
| Honde 2023 [21] | 0 |
| Kamabattula 2026 [19] | 0 |
| Netter 2026 [8] | 0 |

The four-level article appraisal and the named-step count are distinct variables. Four articles had zero named steps in this appraisal although some yielded extracted terms in the blinded round; Ceccaroni 2024 [13], for example, had an appraisal count of zero but 12 and 16 extracted terms, because its steps are narrated rather than named in the text. Seven of the nine articles yielded at least one blinded-round term (Table 1).

The appraisal counted steps named as such in the text; the extraction recorded every operative act the reviewers judged to be a step, including steps narrated in a video or listed in a table without being named as steps. The article-level coding file is deposited in the registration.

**S4. Governance: competing interests and adjudication recusals**

**Financial interests.** Paulo Ayroza Ribeiro is Chief Executive Officer and partner of Épico Women’s Health; Helizabet Salomão Abdalla Ayroza Ribeiro is a partner of the same company without operational role. Mauricio S. Abrão and Henrique Abrão are partners of HerHealth. Paulo Ayroza Ribeiro and Mauricio S. Abrão participate in a revenue-share arrangement on any future commercialization of the ontology under development in the parent project. Neither company is an affiliation of the work, funded it or took part in it. Ana Clara Servidoni, Marina Paula Andres and Guilherme Karam declare no competing interests.

**Intellectual interest intrinsic to the design.** The preliminary ontology that the parent project will submit to consensus was written by the principal investigator. Four safeguards fixed a priori apply to the pilot and to Phase 1: the author of the ontology takes part in neither screening nor extraction; the extraction form contains no column mapping terms to the ontology’s labels; the comparison between the literature-derived inventory and the preliminary ontology is performed by a committee member who neither collected the data nor wrote the labels; and the design admits and reports rejection of labels.

**Overlapping authorship and recusals.** Two articles in the pilot sample are co-authored by members of the steering committee: the AAGL 2021 endometriosis classification [9] (Marina Paula Andres, Mauricio S. Abrão) and the lesion-ontology systematic review by Netter et al [8]. (Henrique Abrão). Neither Mauricio S. Abrão nor Henrique Abrão screened, extracted or adjudicated. Marina Paula Andres, the adjudicator of extraction disagreements, is recused from any article she co-authored; adjudication then passes to a committee member who is not an author of that article and took part in neither its screening nor its extraction — Henrique Abrão in the first instance, Mauricio S. Abrão for articles Henrique Abrão co-authored. Guilherme Karam is excluded from the substitution so that his independence as third reviewer of the 20% re-extraction in Phase 1 is preserved. In the pilot, adjudication of the AAGL 2021 classification was reassigned under this rule; no adjudication arose for the Netter et al. review, from which neither reviewer extracted a term. Every substitution is recorded with the article and the reason.

**S6. Chronology of the pilot and data provenance**

Dates are activity dates supplied by the study coordinator: the manual and instructions were sent to the reviewers on 18 August 2026; the blinded extractions were performed between 18 and 31 August and the completed forms received on 31 August; the analyses, the exploratory exercises and the harmonization session took place between 31 August and 1 September; the directed audit on 2 September; the registration was created on 4 September 2026. The analysis reports (ATLAS-EST-06 to -12) were generated between 31 August and 2 September. Exploratory LLM-assisted exercises are excluded from every estimate but informed the later conventions. An em dash indicates that no interpretable Jaccard estimate is reported for that exercise.

**Supplementary Table S6.** Chronology of the pilot

| **Step** | **Date** | **What it was** | **Arms** | **Studies** | **Mean J** | **Disposition of the records** |
| --- | --- | --- | --- | --- | --- | --- |
| 1 — blinded round | 18–31 Aug 2026 | First independent extraction, blind to each other | Reviewer 1; Reviewer 2 | 7 with a defined index; 2 with no term from either reviewer | 0.581 | Initial independent estimates; records locked |
| 2 — LLM-1 | 31 Aug 2026 | LLM-generated extraction audited by R1; stricter named-step criterion | Audited LLM output; R2 | 9 (4 with terms in both arms) | — | Incomparable instruments/units; excluded from principal analyses |
| 3 — LLM-2 | 31 Aug 2026 | Further comparison of audited exploratory output with R2; four matched terms | As in step 2 | 9 | — | Exploratory only; informed conventions |
| Discussion | 31 Aug–1 Sep 2026 | Discussion, adjudication, and convention revision | R1; R2; committee | 7 | — | Three written conventions adopted; decision log and form version deposited (S2) |
| 4 — post-revision | 1 Sep 2026 | Record revision on previously discussed articles | R1 revision; R2 unchanged | 7 | 0.913 | Convergence, not independent reliability; all 29 additions came from R2 |
| 5 — corrected dataset | 1 Sep 2026 | Nine articles represented; the two zero-term articles declared explicitly by both reviewers | Reviewer 1; Reviewer 2 | 9 (7 with a defined index) | 0.913 | Seven defined indices; zero declared by placeholder row for both empty articles |
| 6 — directed audit | 2 Sep 2026 | Post-hoc start-definition audit | Reviewer 1; Reviewer 2 | 83 matched terms in 7 articles | — | 46/83 discordant responses; 45 in one direction (Table 5) |

**S7. Full contingency tables**

Computed from the deposited datasets atlas_extracao.csv (blinded round; 55 exactly matching terms after normalization; six articles with paired study-level attributes) and atlas_extracao_r4.csv (post-harmonization round; 83 matching terms; seven articles). Rows: Reviewer 1; columns: Reviewer 2. The post-harmonization values are convergence after discussion, not independent estimates.

**Blinded round (55 matched terms)**

**Start definition present**

| **R1 \ R2** | **No** | **Yes** | **Total** |
| --- | --- | --- | --- |
| No | 0 | 20 | 20 |
| Yes | 0 | 35 | 35 |
| Total | 0 | 55 | 55 |

Po = 35/55 = 0.636; κ = 0.000 by construction (Reviewer 2 constant). Nominal 95% CI −0.350 to 0.350.

**End definition present**

| **R1 \ R2** | **No** | **Yes** | **Total** |
| --- | --- | --- | --- |
| No | 46 | 0 | 46 |
| Yes | 1 | 8 | 9 |
| Total | 47 | 8 | 55 |

Po = 54/55 = 0.982; κ = 0.930 (95% CI 0.795–1.000).

**Laterality mentioned**

| **R1 \ R2** | **No** | **Yes** | **Total** |
| --- | --- | --- | --- |
| No | 49 | 0 | 49 |
| Yes | 2 | 4 | 6 |
| Total | 51 | 4 | 55 |

Po = 53/55 = 0.964; κ = 0.781 (95% CI 0.483–1.000).

Anatomical structure (set identity after normalization): 48/55 = 0.873.

Study-level fields, six articles (Abrao, Ceccaroni, Chikazawa, Limbachiya, Nicolì, Raimondo): procedure class — both reviewers “deep endometriosis surgery” in 6/6 (κ not estimable, Po = 1.000); access route — laparoscopic 5, robotic 1, identical for both reviewers (κ = 1.000, Po = 1.000). Honde 2023 has no Reviewer 1 row in the blinded round and therefore no paired study-level attributes.

**Post-harmonization round (83 matched terms)**

**Start definition present (form field, before the directed audit)**

| **R1 \ R2** | **No** | **Yes** | **Total** |
| --- | --- | --- | --- |
| No | 0 | 15 | 15 |
| Yes | 0 | 68 | 68 |
| Total | 0 | 83 | 83 |

Po = 68/83 = 0.819; κ = 0.000 by construction. The directed audit of 2 September 2026 (Table 5) re-examined this field and found 37/83 concordant responses.

**End definition present**

| **R1 \ R2** | **No** | **Yes** | **Total** |
| --- | --- | --- | --- |
| No | 73 | 0 | 73 |
| Yes | 0 | 10 | 10 |
| Total | 73 | 10 | 83 |

Po = 1.000; κ = 1.000.

**Laterality mentioned**

| **R1 \ R2** | **No** | **Yes** | **Total** |
| --- | --- | --- | --- |
| No | 79 | 0 | 79 |
| Yes | 0 | 4 | 4 |
| Total | 79 | 4 | 83 |

Po = 1.000; κ = 1.000.

Anatomical structure: 80/83 = 0.964. Study-level fields, seven articles: procedure class identical in 7/7 (κ not estimable); access route laparoscopic 6, robotic 1, identical (κ = 1.000).

**S8. GRRAS checklist**

Guidelines for Reporting Reliability and Agreement Studies (Kottner J et al., J Clin Epidemiol 2011;64:96–106; reference 12 of the main article). Location refers to the submitted manuscript.

| **Item** | **GRRAS recommendation** | **Location** |
| --- | --- | --- |
| 1 | Identify in title or abstract that the study investigates reliability or agreement | Title (“blinded inter-reviewer agreement study”); Abstract; Highlights |
| 2 | Name and describe the diagnostic or measurement device of interest explicitly | Section 2.4; Supplementary S1, S2 |
| 3 | Specify the subject population of interest | Section 2.3; Table 1 (articles, not patients) |
| 4 | Specify the rater population of interest (if applicable) | Section 2.2 (two reviewers, profiles and reading languages) |
| 5 | Describe what is already known about reliability and agreement and provide a rationale for the study | Introduction, paragraphs 1–2 (references 1–5) |
| 6 | Explain how the sample size was chosen; state the determined number of raters, subjects/objects and replicate observations | Section 2.3 (purposive stress-test sample of 11; rationale); Section 4.2 (limitations of size) |
| 7 | Describe the sampling method | Section 2.3 (purposive selection by the steering committee; not a systematic search) |
| 8 | Describe the measurement/rating process (time interval between repeated measurements, availability of clinical information, blinding) | Sections 2.2, 2.5; Supplementary Table S6 (blinding between reviewers; no translation aid; harmonization session; unchanged Reviewer 2 records) |
| 9 | State whether measurements/ratings were conducted independently | Section 2.5 (blinded round independent; post-harmonization round not independent) |
| 10 | Describe the statistical analysis | Sections 2.6, 2.8 |
| 11 | State the actual number of raters and subjects/objects included and the number of replicate observations conducted | Section 3.1; Table 1; Table 2 (n per estimate) |
| 12 | Describe the sample characteristics of raters and subjects (e.g. training, experience) | Section 2.2; Table 1 |
| 13 | Report estimates of reliability and agreement including measures of statistical uncertainty | Sections 3.2–3.5; Tables 2–6; Figure 1; Supplementary S7 (nominal CIs for κ; no CI for Jaccard, with reasons) |
| 14 | Discuss the practical relevance of results | Sections 4, 4.1 (implications for evidence synthesis), 4.3 |
| 15 | Discuss the limitations of the study | Section 4.2 |
| 16 | Report other/further findings, if applicable | Section 3.4 (post-hoc audit of the start-definition field); Supplementary S3, S9 |

**S9. Computational integrity audit of the two record-integrity defects**

Two defects were found and corrected during analysis; both are reported in section 2.8 of the main article, with the before-and-after outputs deposited. The corrected analysis script (version 1.1) and its output are in the associated public project (https://osf.io/vca2s/, folder “Post-registration corrections”).

**Supplementary Table S9.** First defect — locale-dependent matching

| **Item** | **Before correction** | **After correction** |
| --- | --- | --- |
| Studies expected in the paired analysis | 7 | 7 |
| Studies actually analyzed | 6 | 7 |
| Study silently dropped | Nicolì 2024 [18] | none |
| Jaccard index of the dropped study | 0.231; second-lowest (Honde: 0.000) | 0.231, included |
| Mean Jaccard index, paired set | 0.639 | 0.581 |
| Cause | Study names matched as literals; under a C/POSIX locale a non-ASCII name failed to match its UTF-8 record | Explicit UTF-8 encoding and a locale guard in all scripts |
| Integrity test | Absent | Assertions on the expected number of studies and term rows, uniqueness of study identifiers, and failure on any dropped record |

Omitting Nicolì increased the seven-article mean from 0.581 to 0.639 because its index was below the mean; it was not the lowest-performing article (Honde: 0.000). The before-and-after datasets and outputs are deposited with article-level joins and fail-fast identity checks.

Second defect — placeholder row counted as a term. Reviewer 1 declared a zero-term article (Netter 2026 [8]) with a single placeholder row whose term field read ‘NR’ (not reported), following the form’s instruction to declare zero explicitly rather than leave the article blank. The first analysis treated that row as a term, giving Netter a defined index of zero, an eight-article mean of 0.508, a pooled index of 55/93 and a record count of 148. Because both reviewers extracted no term, the article has an undefined index and is excluded under the prespecified rule; the corrected values are the seven-article mean of 0.581, the pooled index of 55/92 and 147 records. The corrected scripts treat the placeholder marker as a declared zero. The outputs before and after both corrections are deposited in the registration (https://doi.org/10.17605/OSF.IO/GMXZ5); the corrected script (version 1.1) and its output are in the associated public project (https://osf.io/vca2s/).

**S10. The four extraction conventions and the evidence-traceability rules**

Adopted at the harmonization session of 31 August–1 September 2026 (conventions 1–3) and after the analysis of the French-language survey (convention 4); incorporated in the extraction manual (Appendix B-1 v3.0) and the revised form (Appendix B v3.0) deposited in the registration. They govern the Phase 1 extraction and the re-extraction of the nine pilot articles.

**Convention 1 — What counts as a step.** Extract the operative act that the authors name as a step, phase or stage, including acts presented in numbered lists, section headings, table rows and figure or video captions. Do not extract a therapeutic modality, technique, procedure or diagnosis that the article names without presenting it as part of an operative sequence. A four-question test in the manual decides borderline cases, with a closed table of fifteen decided examples.

**Convention 2 — Extent of the term.** Where a published label exists, the term is the full text of that label up to the first full stop, semicolon or colon. Where there is no label, the term is the shortest noun phrase the authors use for the act. Qualifiers of instrument, size or position are part of the term only when they are part of the label.

**Convention 3 — Scope of reading.** The reviewer reads the abstract, the body text, the tables, the figure and video captions and the supplementary material of the article, and nothing beyond that. Each term carries a locator (section and page, or timestamp) precise enough to take the adjudicator straight to the source.

**Convention 4 — Source in a language the reviewer does not read fluently.** The reviewer may use a translation aid for comprehension, and records which aid was used. The term is transcribed in the original language, never in translation; the translated passage is not the datum. A passage the reviewer cannot resolve with the aid is flagged for verification by a reader of the language, and is not coded as absence of a term. The use of an aid and the reviewer’s declared reading languages are recorded per article.

**Evidence-traceability rules.** The former single field “Does the article state when the step begins?” was split into two: (a) an operational start definition stated in the published text, answered yes only with the verbatim quotation and its locator; and (b) a step demonstrated on video, answered separately with the timestamp, and never counted as a textual definition. The same applies to the end definition. “Not assessed” is recorded separately from “absent”. An article that yields no term receives one row declaring zero, with a fixed marker in the term field and a justification, so that an empty article is never silently absent from the dataset; the analysis scripts treat that marker as a declared zero, not as a term.
